# Determinants of Emergency Contraceptive Use Among Adolescent Girls Aged 15–19 Years in West Asembo Ward, Rarieda Sub-County, Siaya County, Kenya. A cross sectional study

**DOI:** 10.64898/2026.10.29.26366853

**Authors:** George O. Ang’ang’o, Daniel Onguru, Sidney Ogolla, Iwaret Otiti

## Abstract

Despite high uptake of conventional contraceptive services, adolescent pregnancies remain high in West Asembo ward, Siaya County, Kenya. This study determined the utilization, factors associated with use, and perceptions towards emergency contraception (EC) among adolescent girls in this region. A cross-sectional study design using a mixed-methods approach was employed. Quantitatively, 400 female adolescents aged 15–19 years were randomly sampled using stratified sampling across five sub-locations. Data were collected via interviewer-administered electronic questionnaires. Qualitatively, ten purposively selected Key Informant Interviews were conducted with healthcare and pharmaceutical providers, alongside four Focus Group Discussions with adolescents in sub-locations representing the highest and lowest teenage pregnancy rates. Quantitative data were analyzed using descriptive and inferential statistics (α=0.05) in SPSS v.29, while qualitative data underwent thematic analysis in NVivo 14 before triangulation. Results showed that 34.8% of the respondents had ever used emergency contraception. In the bivariate analysis, being aged 18–19 years significantly reduced the likelihood of EC use compared to those aged 15–17 years (COR: 0.32, 95% CI: 0.21–0.50, p < 0.0001). Lower primary education was associated with a lower likelihood of EC use compared to no formal education (COR: 0.04, 95% CI: 0.003–0.49, p = 0.012). Crucially, adolescent girls who were single and living alone were significantly more likely to use ECs than single counterparts living with parents or guardians (COR: 5.23, 95% CI: 2.86–9.56, p < 0.0001). In conclusion, emergency contraceptive use among adolescent girls in West Asembo Ward is relatively low. The findings underscore the need for a comprehensive, adolescent-centered approach involving health facilities, schools, community health promoters, and private pharmacies to empower adolescents with accurate information while ensuring confidential, accessible EC services coupled with reproductive health counseling.

## METHODOLOGY

### Study setting and design

The study was conducted in West Asembo Ward, Rarieda Sub-County, Siaya County, Kenya. The ward covers 101.0 km² and comprises five sub-locations: Siger, Mahaya, Akom, Memba, and Nyagoko. According to the 2019 Kenya Population and Housing Census, the ward had a population of 33,102, with subsistence crop and livestock farming as the predominant economic activity. The study employed a convergent parallel mixed-methods cross-sectional design, involving quantitative questionnaires and qualitative Key Informant Interviews (KIIs) and Focus Group Discussions (FGDs).

### Study population and sampling

The target population comprised adolescent girls aged 15–19 years who had resided continuously in West Asembo Ward for at least 36 months. The quantitative sample size was determined using Cochran’s formula and adjusted for the finite population, resulting in a minimum sample of 361; a 10% allowance for non-response yielded a target sample of 400 adolescents. Stratified random sampling was used to select participants across the five sub-locations, with sampling frames developed from household registers in collaboration with Community Health Promoters.

For the qualitative component, purposive sampling was used to select 10 key informants, comprising five healthcare providers and five pharmaceutical technologists. Four FGDs, each comprising 8–10 adolescent girls, were conducted in Nyagoko and Memba sub-locations, selected to represent areas with relatively high and low adolescent pregnancy burden, respectively.

### Data collection and analysis

Data was collected from 10^th^ June 2024 to 30^th^ Dec 2024. Quantitative data were collected using interviewer-administered structured questionnaires and captured electronically through Google Forms. Five trained field assistants participated in data collection, with each assigned to one sub-location. Data were reviewed daily for completeness and consistency. Qualitative data were collected using semi-structured KII and FGD guides. Interviews and discussions were audio-recorded with participants’ permission, transcribed, anonymized, and thematically analyzed using NVivo version 14.

Quantitative data were cleaned using SPSS Builder version 4.0 and analyzed using SPSS version 29. Descriptive statistics included frequencies, percentages, means, and standard deviations. Associations between emergency contraceptive use and categorical variables were assessed using chi-square tests. Variables significant at *p* < 0.05 in bivariate analysis were entered into a multivariable binary logistic regression model. Results were presented as crude and adjusted odds ratios (COR and AOR) with 95% confidence intervals, with statistical significance set at *p* < 0.05. Qualitative findings were triangulated with quantitative results to enhance interpretation.

### Ethical considerations

Ethical approval was obtained from the University of Eastern Africa, Baraton Ethics Review Committee (UEAB/REC/30/04/2024), while authorization was obtained from the JOOUST Board of Postgraduate Studies and a research license from NACOSTI. Written informed consent was obtained from participants aged 18–19 years, while parental/guardian consent and participant assent were obtained for those below 18 years. Confidentiality, anonymity, and voluntary participation were maintained throughout the study.

## RESULTS

A total of 400 adolescent girls were recruited for the study. One questionnaire was excluded due to data incompleteness, yielding a final sample of 399 respondents (99.75% response rate). The quantitative survey revealed an emergency contraception (EC) prevalence rate of 34.8% (= 139) among the respondents. These quantitative data were triangulated with qualitative themes derived from four Focus Group Discussions (FGDs) with adolescent cohorts and ten Key Informant Interviews (KIIs) with local healthcare providers and pharmaceutical technologists.

### Sociodemographic Characteristics of Respondents

The baseline sociodemographic profile of the analytical sample is summarized in Table 1. The majority of the respondents were aged between 18 and 19 years (236, 59.1), with an overall cohort mean age of 17.14 ± 1.27 years.

Regarding marital status and domestic living arrangements, the majority of the participants were single and residing with parents or guardians (309, 77.4%). A distinct subset comprised single adolescents living independently (58, 14.5%), while the remainder were married and cohabiting with a spouse (25, 6.3%) or married but living away from their partner (7, 1.8%).

**Table 3:**
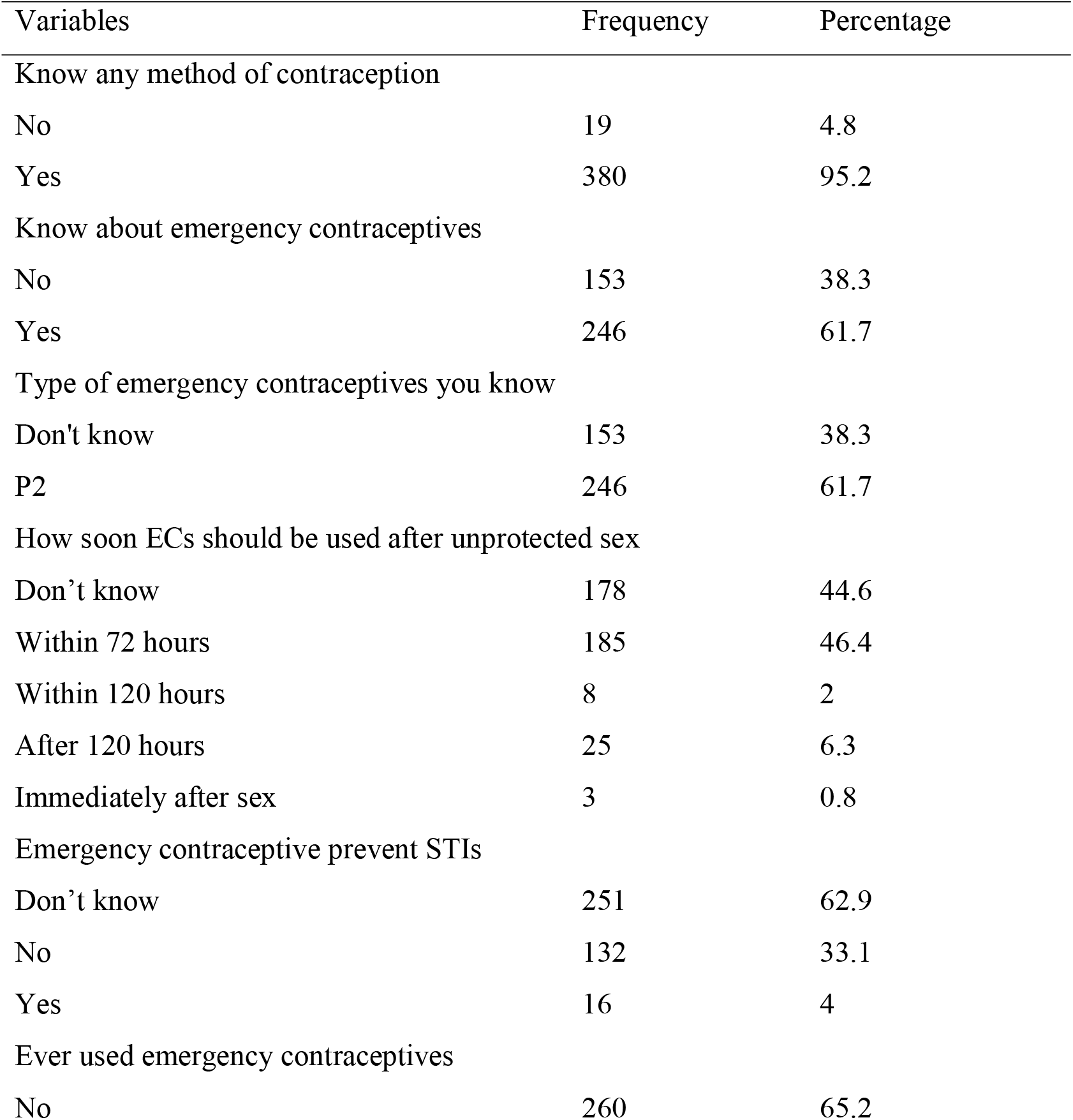
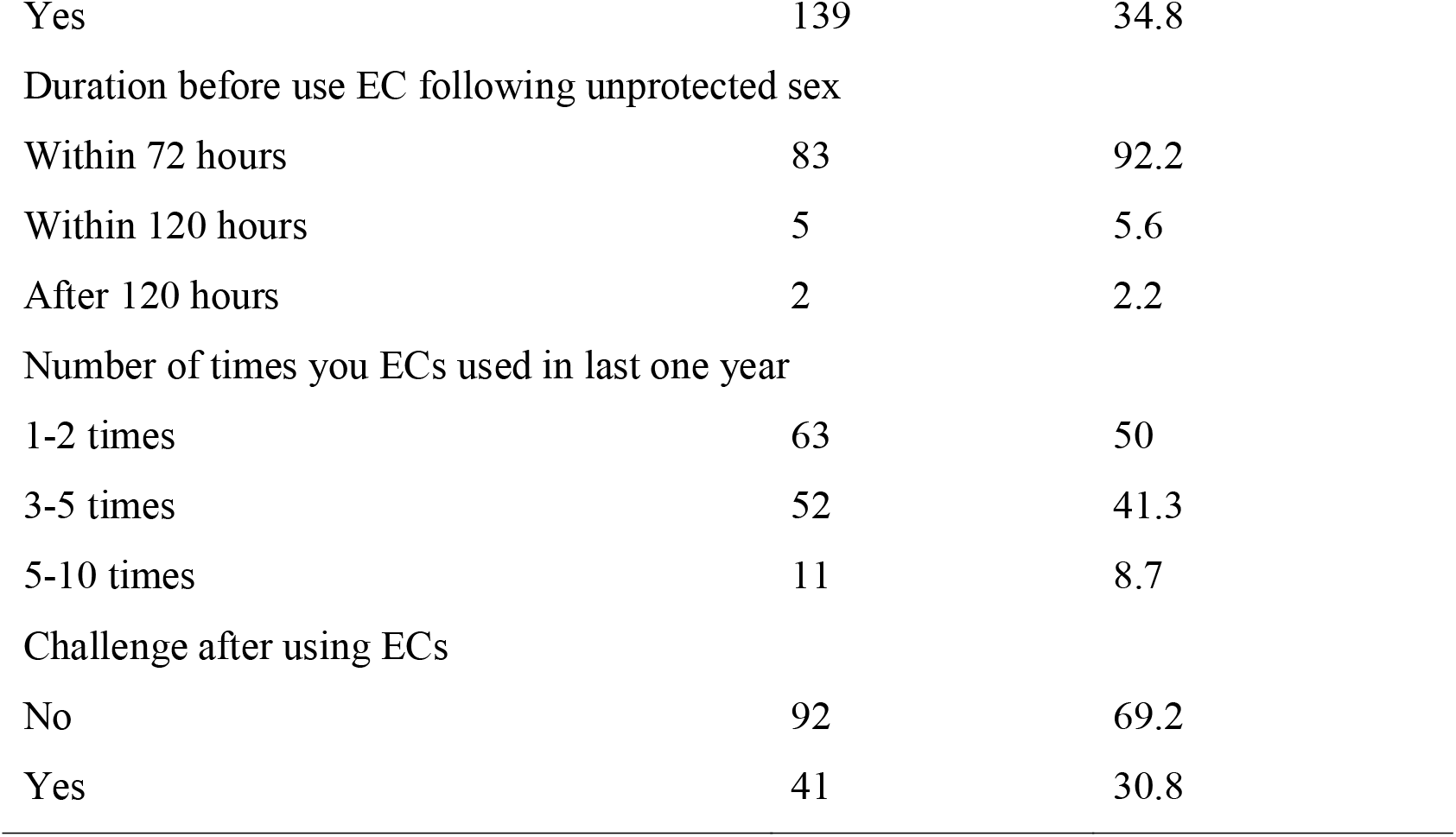
Knowledge of emergency contraceptives.

The dominant primary occupation was school enrollment, with 67.4% (269) of the cohort classified as students. Regarding physiological reproductive milestones, the onset of menarche was most frequently documented between the ages of 11 and 13 years, accounting for 77.7% (310) of the study population.

### Reproductive Characteristics and Emergency Contraception Uptake

Approximately one-third of the participants (139, 34.8%) reported having used ECs, with higher usage among those with a history of pregnancy (57, 40.4%) compared to those without (82, 31.8%). However, pregnancy history did not significantly affect EC use (p=0.083). Adolescents with children significantly showed higher EC usage rates, with 19(48.7%) of those with two children, 37(41.1%) of those with one child, and 82 (31.3%) of those without children reporting EC use (p=0.044). However, there was no significant correlation found between unintended pregnancy (p=0.095) and history of abortion (p=0.071) and the use of ECs (Table 4.2).

**Table 4.1:**
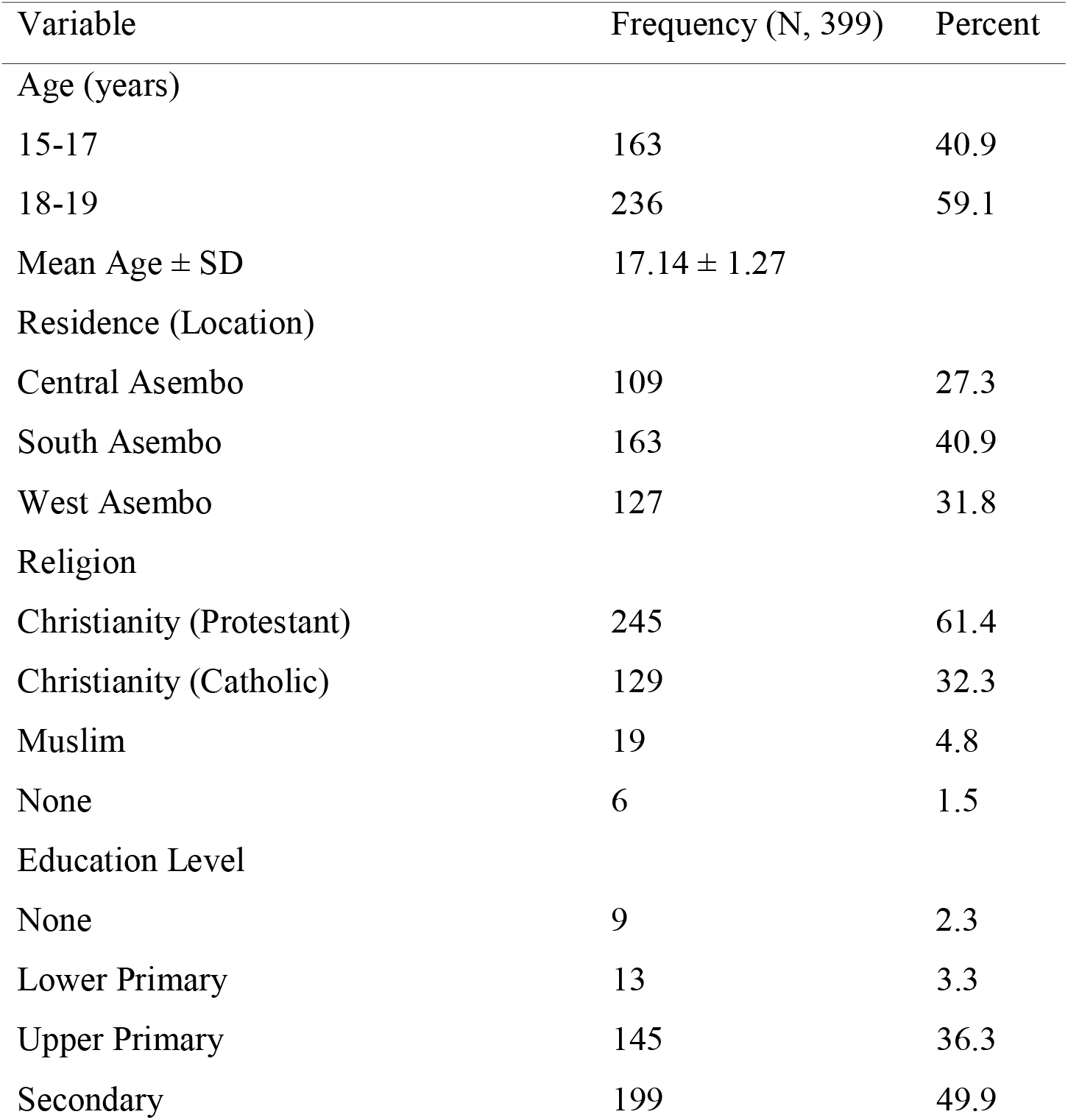
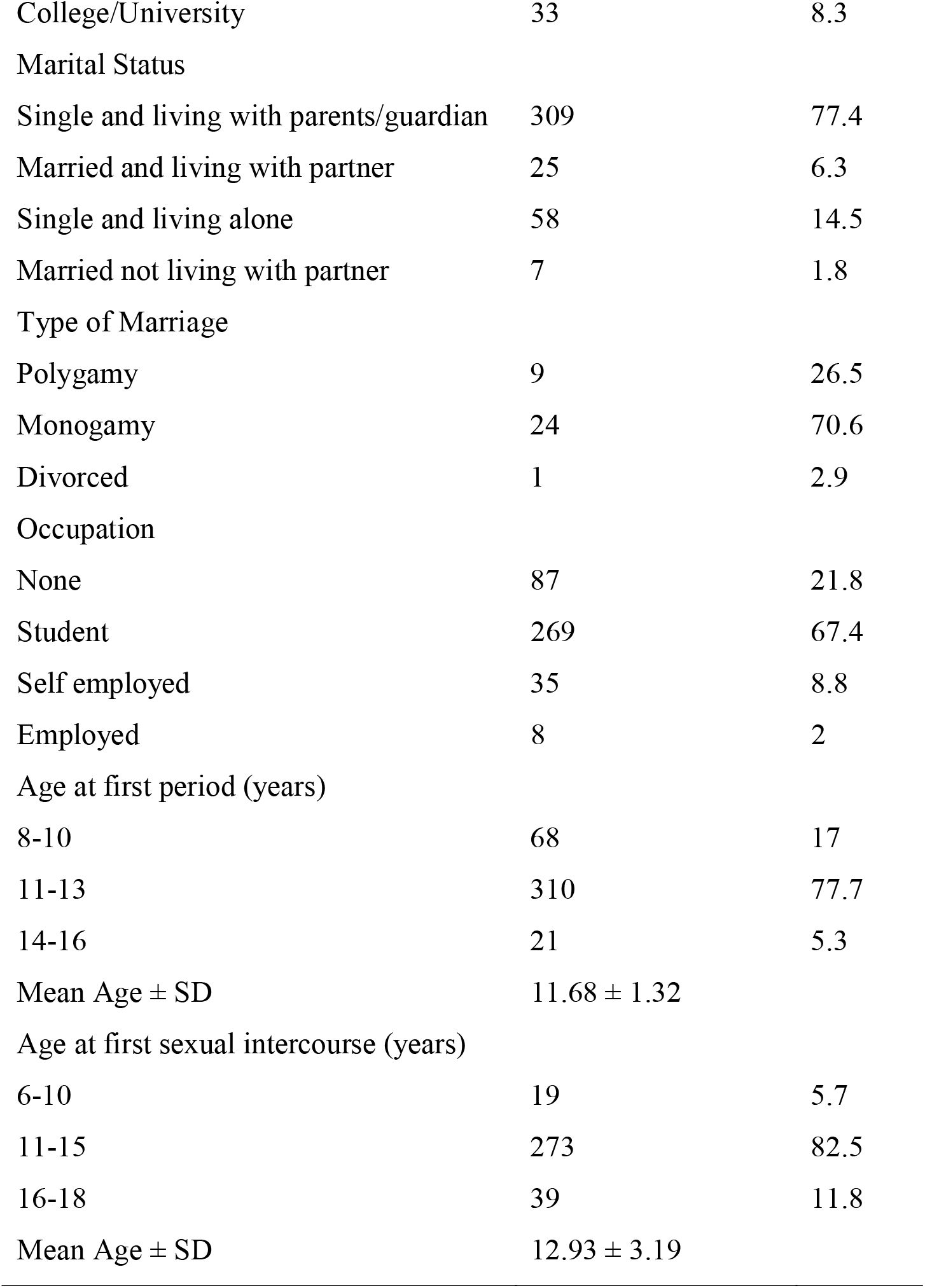
Sociodemographic characteristics of the respondents.

| Variable | Frequency (N, 399) | Percent |
| --- | --- | --- |
| Age (years) |  |  |
| 15-17 | 163 | 40.9 |
| 18-19 | 236 | 59.1 |
| Mean Age ± SD | $17.14 \pm 1.27$ | |
| Residence (Location) |  |  |
| Central Asembo | 109 | 27.3 |
| South Asembo | 163 | 40.9 |
| West Asembo | 127 | 31.8 |
| Religion |  |  |
| Christianity (Protestant) | 245 | 61.4 |
| Christianity (Catholic) | 129 | 32.3 |
| Muslim | 19 | 4.8 |
| None | 6 | 1.5 |
| Education Level |  |  |
| None | 9 | 2.3 |
| Lower Primary | 13 | 3.3 |
| Upper Primary | 145 | 36.3 |
| Secondary | 199 | 49.9 |
| College/University | 33 | 8.3 |
| Marital Status |  |  |
| Single and living with parents/guardian | 309 | 77.4 |
| Married and living with partner | 25 | 6.3 |
| Single and living alone | 58 | 14.5 |
| Married not living with partner | 7 | 1.8 |
| Type of Marriage |  |  |
| Polygamy | 9 | 26.5 |
| Monogamy | 24 | 70.6 |
| Divorced | 1 | 2.9 |
| Occupation |  |  |
| None | 87 | 21.8 |
| Student | 269 | 67.4 |
| Self employed | 35 | 8.8 |
| Employed | 8 | 2 |
| Age at first period (years) |  |  |
| 8-10 | 68 | 17 |
| 11-13 | 310 | 77.7 |
| 14-16 | 21 | 5.3 |
| Mean Age ± SD | 11.68 ± 1.32 | |
| Age at first sexual intercourse (years) |  |  |
| 6-10 | 19 | 5.7 |
| 11-15 | 273 | 82.5 |
| 16-18 | 39 | 11.8 |
| Mean Age ± SD | 12.93 ± 3.19 | |

**Table 4.2:**
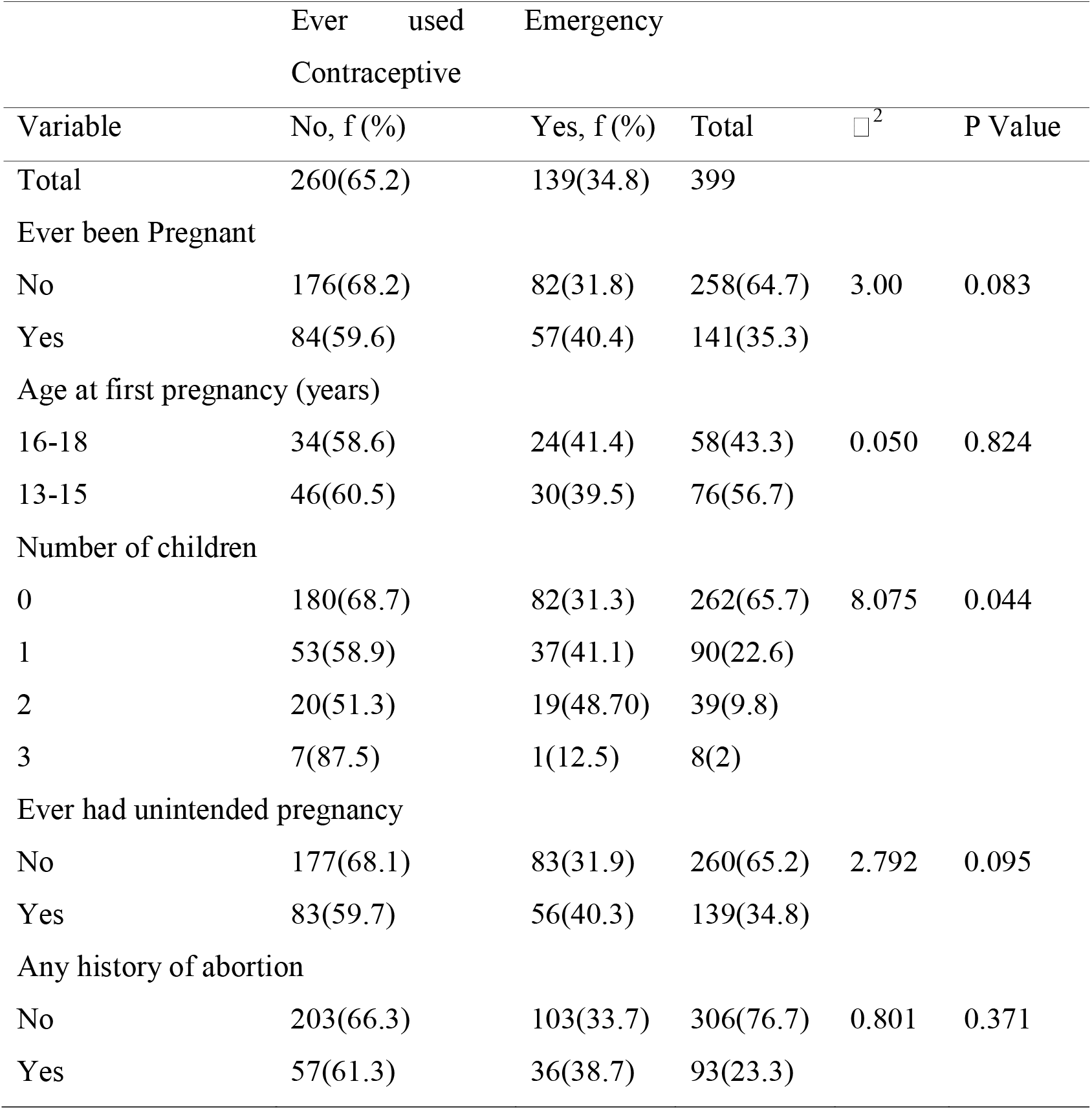
Reproductive characteristics and emergency contraceptive uptake.

The KIIs with chemists supported these findings, indicating a high demand for ECs among adolescents and college students, as reflected in the sentiments below:

> “The teenagers of ages between 18-22 years are the primary users of P2” (KC 05) “Young ladies ages between 15-19 years of age are the majority P2 clients” (KC04)

The chemists noted rapid stock turnover, suggesting high utilization rates, though misuse was a concern, with some adolescents reportedly using ECs as routine contraception rather than for emergencies. One chemist attendant KC01 noted that *“emergency pill goes very fast; its stock cannot take even two weeks.”*

However, misuse of ECs was noted by the same attendant:

> “I see people using it like a normal family planning method. They don’t use it on emergency cases” A chemist attendant KC03 further mentioned, “I was once told that P2 shouldn’t be taken frequently but nowadays I see some people taking it anytime they have sex.”

### Sources of Emergency Contraceptives reported by respondents

Retail commercial pharmacies and local chemists serve as the primary source of emergency contraception among adolescent girls, supplying up to 73% of the demographic either exclusively or alongside peer networks. Formal public and private healthcare facilities account for a minimal proportion of distribution at 3% each, while 18% of participants report a complete lack of awareness regarding where to legally acquire the commodities.

**Figure 1:**
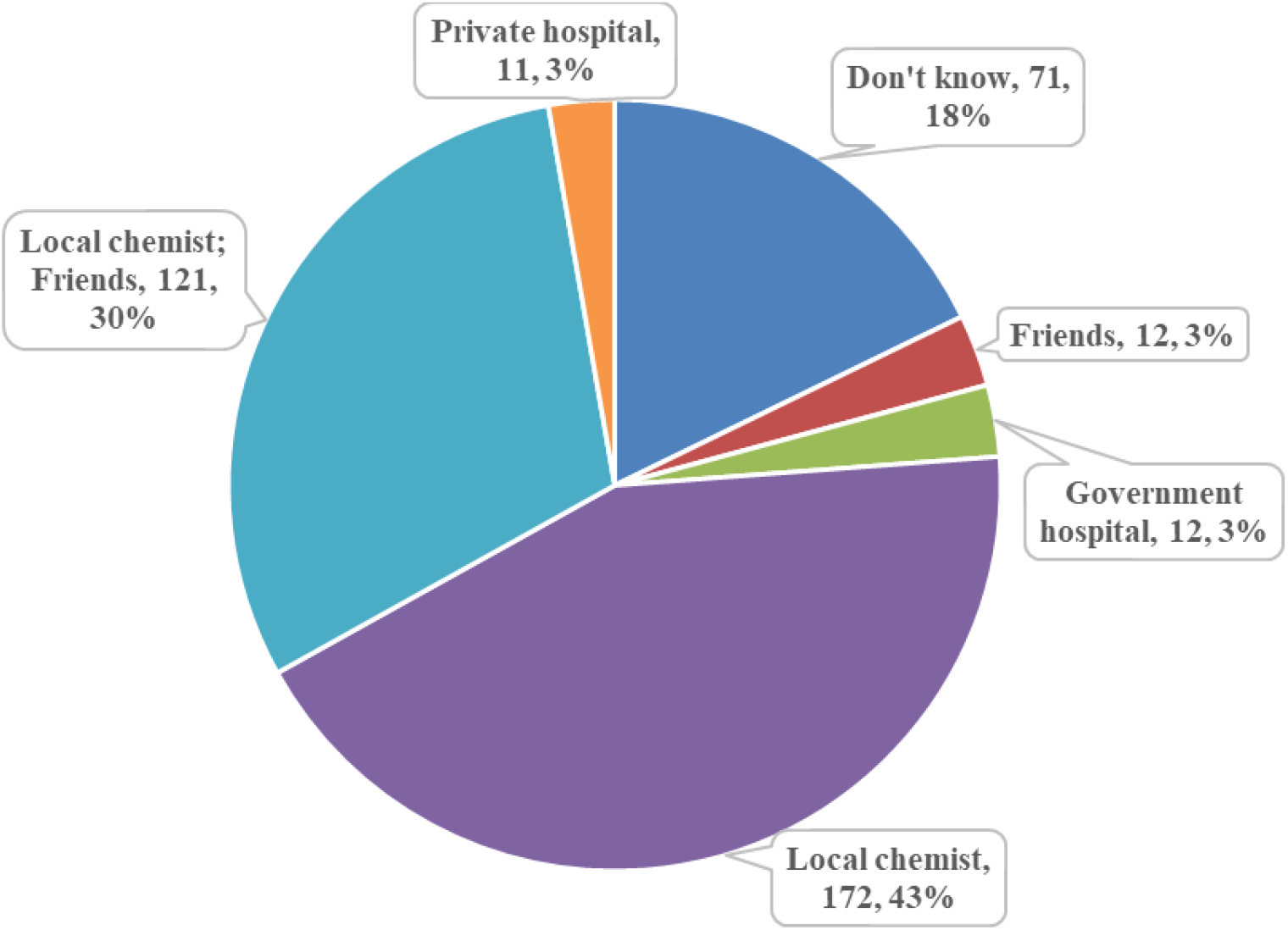
Sources of Emergency Contraceptives reported by respondents

Focus Group Discussions (FGDs) with adolescent cohorts strongly corroborated the quantitative preference for the commercial retail sector over public infrastructure. Participants consistently identified private retail chemists, male partners, and peer networks as their primary sources.

Formal public dispensaries and health centers were rarely cited as viable options. Adolescents noted that male sexual partners frequently took responsibility for purchasing and delivering the commodities directly from chemists.

Key Informant Interviews (KIIs) with facility-based nursing and clinical officers revealed severe, systemic supply-chain failures within the public healthcare tier. Public clinics predominantly stocked long-acting reversible contraceptives (LARCs) and daily oral pills, while emergency contraceptives (ECs) were systematically unavailable due to erratic secondary supply links from sub-county referral hubs.

Chronic public sector stock-outs actively shifted healthcare-seeking behavior away from public clinics toward local commercial pharmacies, where product availability was guaranteed. This chronic unavailability fostered a long-term community perception that public health facilities did not stock ECs. Consequently, adolescent clinic attendance remained heavily suppressed even during periods when the commodity was successfully restocked.

Facility-level demonstrated exceptionally low public consumption rates and prolonged shelf stagnation for public sector EC allocations. Multiple institutional providers reported having issued fewer than two units over a three-to-four-month period despite holding active, restocked inventories.

Both clinical providers and retail pharmaceutical technologists reported clear temporal spikes in EC demand. Peak utilization periods strongly correlated with academic calendars and school holidays, specifically during the months of April, August, and December. Commercial dispensers confirmed that these seasonal influxes rapidly accelerated retail inventory turnover rates, straining local supply capacities and exhausting shelf stocks within two weeks.

### Knowledge and Perception of emergency contraceptives

General awareness of modern contraceptive methods was documented in 380 out of 399 participants (95.2%). However, specific knowledge regarding emergency contraception (EC) was limited. While 246 (61.7%) adolescent girls reported prior awareness of EC, 178 (44.6%) respondents did not know the correct therapeutic window necessary to ensure efficacy.

Sociomedical misconceptions were prevalent among the respondents. A substantial proportion erroneously believed that EC consumption leads to infertility. Additionally, 16 respondents (4.0%) incorrectly stated that EC methods gives protection against sexually transmitted infections (STIs).

Among adolescent girls who reported prior use of EC, 83 of 90 users (92.2%) adhered to recommended clinical timing protocols by administering the regimen within 72 hours post-coitus. Concerning the frequency of administration over the preceding 12 months, 63 out of 126 active users (50.0%) reported using EC between one and two times with 41 out of 133 users (30.8%) reporting post-administration side effects and complications.

Focus Group Discussions (FGDs) with adolescent girls revealed that despite universal community familiarity with the local commercial brand name “P2”, deep-seated deficits in knowledge and significant reproductive health misconceptions remained prevalent. A dominant thematic concern across the discussion cohorts was the conviction that frequent or repeated use of emergency contraception (EC) would cause harm like spontaneous abortion, secondary infertility, and barrenness. These functional distortions regarding safety thresholds and long-term clinical sequelae were expressed uniformly across both geographic strata, underscoring systemic community anxiety surrounding hormonal contraception.

Representative narratives from the participants highlighted these pervasive anxieties:

> “They should be taken once a year within 72 hours after sex. If taken more than thrice a year, it can lead to several complications like abortion, irregular periods, and barrenness.” (FGD, Participant M01, Memba Sub-location)

Another participant echoed similar concerns stating annual usage limit and permanent reproductive damage:

> “It is said that if you take more than twice in a year, it can lead to infertility and barrenness.”

(FGD, Participant N05, Nyagoko Sub-location)

### Other contraceptive methods known to the respondents

The baseline assessment of contraceptive awareness among the participants demonstrated that long-acting reversible contraceptives (LARCs) were the most frequently recognized modalities.

Implants were the most widely identified method (75/399), followed by condoms (67/399) and injectable contraceptives (61/399).

Dual-method awareness was less frequently reported by the respondents. A combination of condoms and implants was reported by 43 participants. In contrast, combinations involving oral contraceptives (25/399) and injectable (21/399) represented the least commonly known contraceptive methods within the study population.

**Figure 2:**
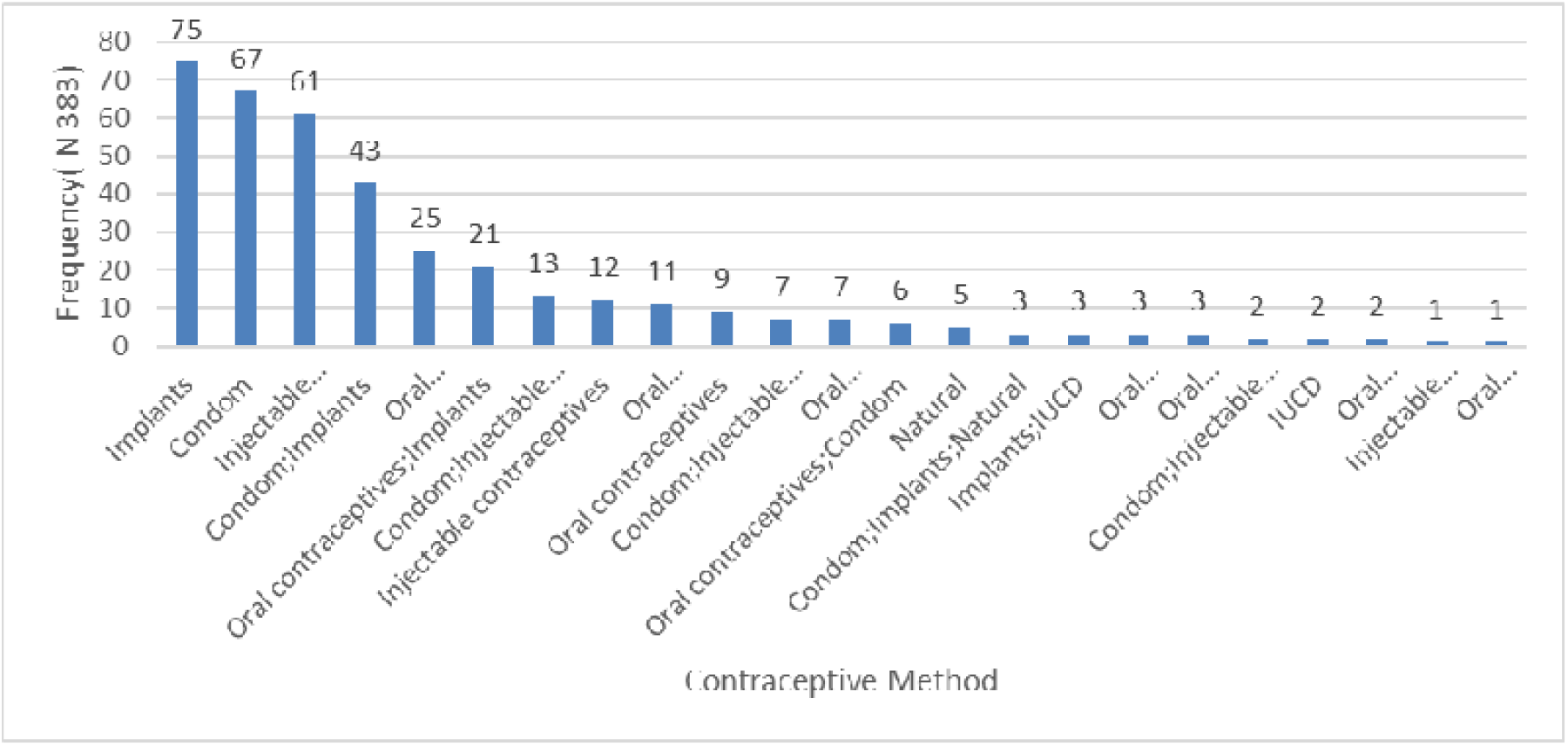
Other Contraceptive Methods known to Respondents

### Situations associated with emergency contraceptive use

Quantitative analysis revealed that sexual assault was the most frequently cited indication for emergency contraception (EC) use, reported by 181 out of 399 respondents (45.4%). This was followed by dual indications involving either sexual assault or condom failure (57/399, 14.3%), and isolated condom failure (53/399, 13.3%).

These empirical findings were corroborated by qualitative data from Focus Group Discussions (FGDs) and Key Informant Interviews (KIIs), which demonstrated that EC is primarily conceptualized within the community as a reactive intervention for forced or unplanned sexual encounters. During the FGD sessions, participants highlighted this crisis-driven utilization pattern:

> *“They are things like rape.”* (FGD, Participant N07, Nyagoko Sub-location)

This systemic reliance on EC for clinical crisis management was further confirmed during a KII at a local dispensary. A frontline healthcare provider observed that adolescents formed the primary demographic seeking EC following unprotected or coerced sex:

> “Mostly youths and teenagers are our clients within the age of 16 to 24 years; then also for rape cases we usually give them.” (KII, Healthcare Professional 05, Local Dispensary)

**Figure 3:**
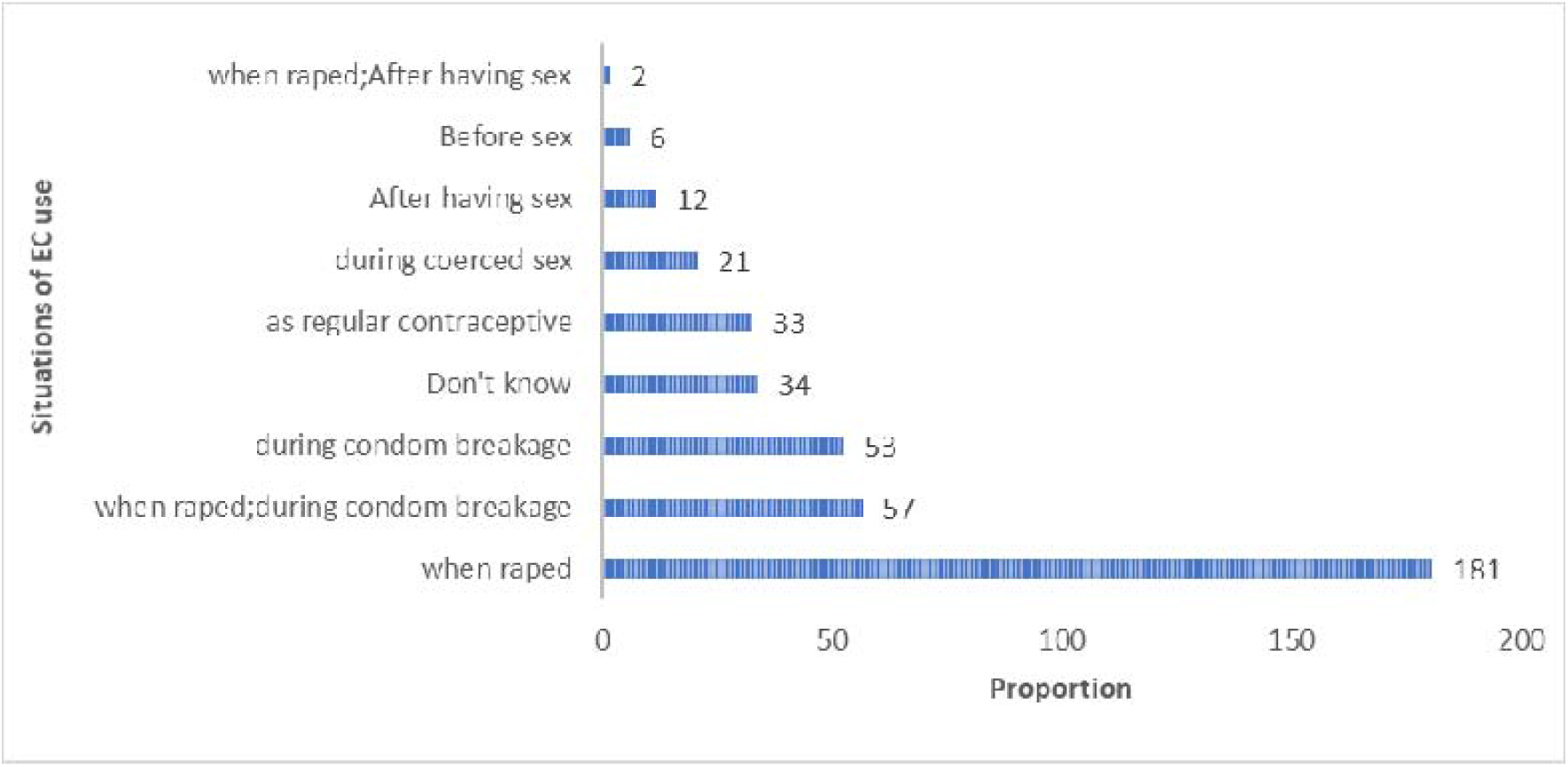
Situations reported as leading to Emergency Contraceptive Use

### Perceived side effects of emergency contraceptives

Descriptive analysis demonstrated that irregular period was the most frequently perceived side effect of emergency contraception (EC), cited by 71 out of 399 respondents (17.8%). This was followed by fears concerning spontaneous abortion or infertility, which were reported by 67 out of 399 participants (16.8%) (Figure 4).

**Figure 4:**
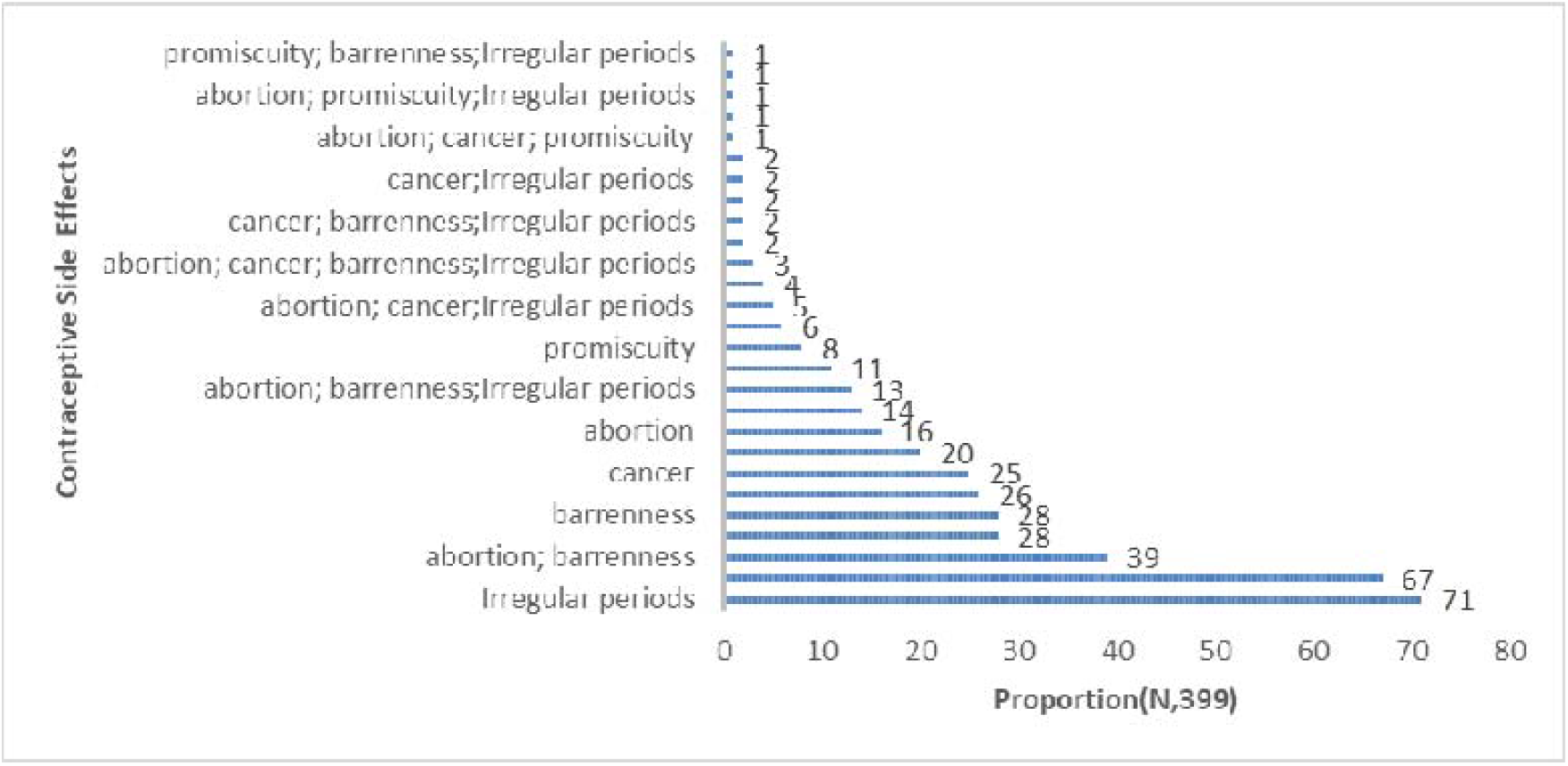
Perceived Side effects of emergency contraceptives

Focus Group Discussion (FGD) strongly corroborated these quantitative metrics, with participants consistently attributing severe and permanent reproductive effects to frequent EC use. Narratives from the sessions highlighted systemic anxieties regarding uterine damage and permanent sterility:

> “It’s said that it weakens the womb and leads to barrenness.” (FGD, Participant M09, Memba Sub-location)

In addition to anxieties surrounding sub-fertility, participants articulated specific fears regarding uterine cancer and complete lose of periods from hormonal contraceptive use:

> “Others also said that it can lead to cancer of the uterus and can make one completely lose her periods.” (FGD, Participant M07, Memba Sub-location)

Chemists emphasised the need for targeted community education to correct such misconceptions:

> “Let the adolescent girls have the correct information concerning these emergency pills… whatever they have from this community demoralizes them from using these P2” (KC05)

### Sociodemographic determinants of Emergency Contraceptive use

In bivariate analysis, age, education level, marital/living status, and occupation were significantly associated with EC use. Adolescents aged 18–19 years were less likely to have used EC than those aged 15–17 years (COR 0.32, 95% CI 0.21–0.50, p < 0.0001). Compared with respondents with no formal education, those with lower primary education had markedly lower odds of EC use (COR 0.04, 95% CI 0.003–0.49, p = 0.012), as did those with upper primary education (COR 0.08, 95% CI 0.01–0.36, p = 0.001). Adolescents who were single and living alone had substantially higher odds of EC use than those single and living with parents or guardians (COR 5.23, 95% CI 2.86–9.56, p < 0.0001). Students had lower odds of use than those with no occupation (COR 0.55, 95% CI 0.33–0.91, p = 0.021). Religion was not significantly associated with EC use.

In the multivariable model, lower primary education (AOR 0.06, 95% CI 0.004–0.92, p = 0.044) and living alone while single (AOR 3.61, 95% CI 1.63–8.03, p = 0.002) remained independent predictors of EC use, while age and occupation were attenuated to non-significance (Table 4).

### Reproductive and knowledge - based determinants of emergency contraceptive use

Number of children was associated with EC use in bivariate analysis: adolescents with two children had over twice the odds of EC use of those with none (OR 2.08, 95% CI 1.05–4.11, p = 0.034), although this association did not persist after adjustment. Knowledge-related factors showed the strongest associations with EC use in bivariate analysis: awareness of any contraceptive method (OR 10.6, 95% CI 1.35–77.72, p = 0.024), awareness of EC specifically (OR 10.41, 95% CI 5.69–19.05, p < 0.0001), and knowledge of “P2” by name (OR 11.3, 95% CI 6.07–21.04, p < 0.0001) were all strongly associated with higher odds of use, though these associations were not independently significant after adjustment, likely reflecting collinearity among the knowledge variables. Conversely, the mistaken belief that EC remained effective beyond 120 hours after intercourse was independently associated with lower odds of EC use in both bivariate (OR 0.35, 95% CI 0.14–0.85, p = 0.021) and multivariable analysis (AOR 0.34, 95% CI 0.13–0.85, p = 0.021) (Table 5).

**Table 5.**
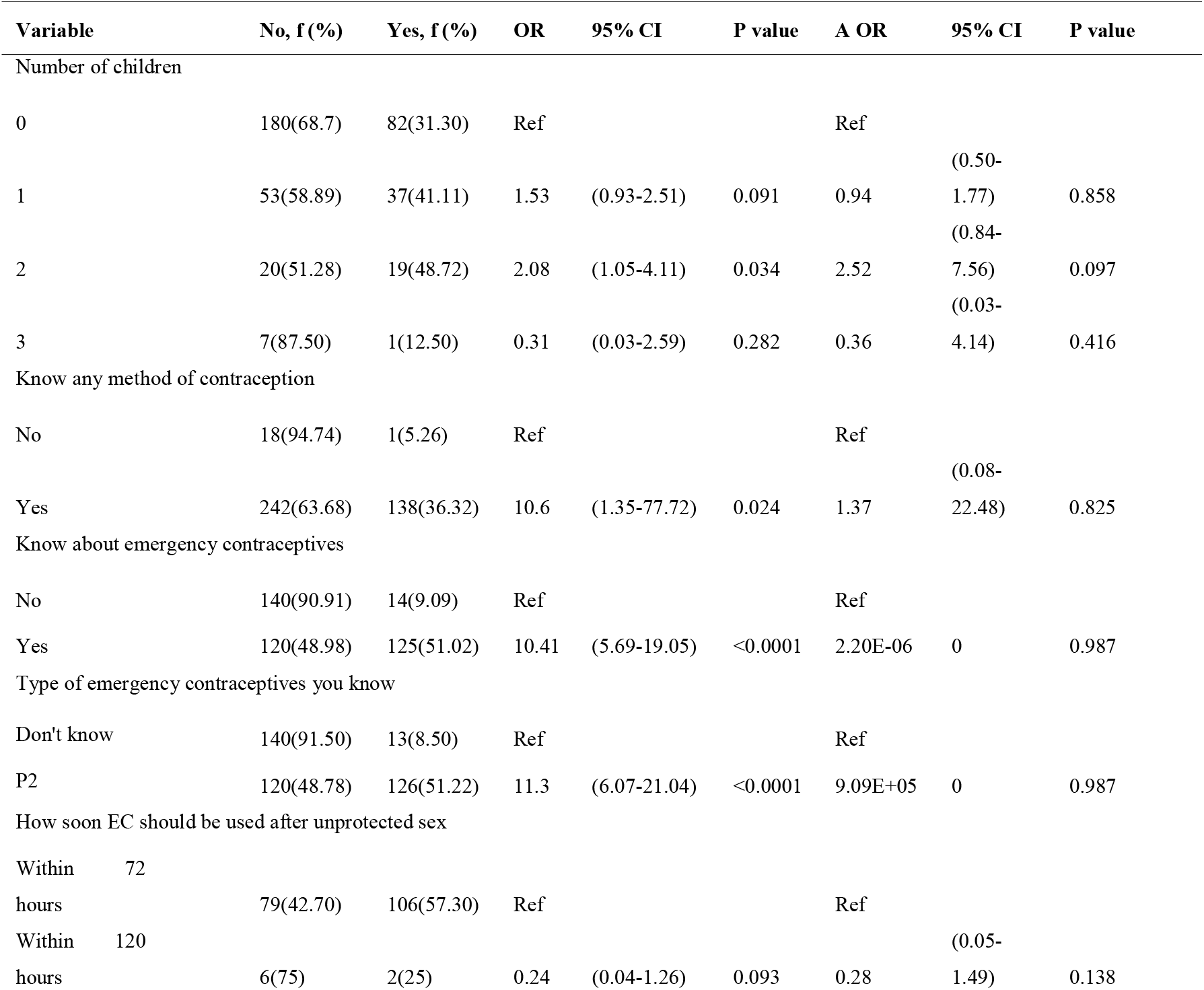
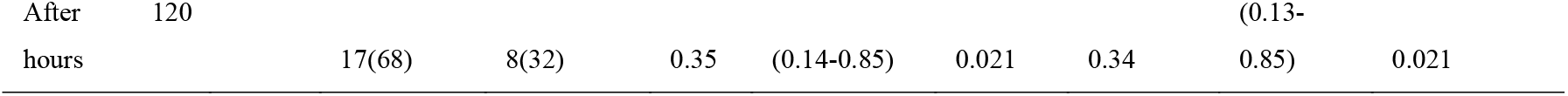
Reproductive and knowledge-based determinants of emergency contraceptive use.

**Table 6.** Sociodemographic determinants of emergency contraceptive use.

| Variables | No, f (%) | Yes, f (%) | COR | 95% CI | P value | AOR | 95% CI | P value |
| --- | --- | --- | --- | --- | --- | --- | --- | --- |
| Age (years) |  |  |  |  |  |  |  |  |
| 15-17 | 82(50.31) | 81(49.69) | Ref |  |  | Ref |  |  |
| 18-19 | 178(75.42) | 58(24.58) | 0.32 | (0.21-0.50) | <0.0001 | 0.63 | (0.36-1.12) | 0.122 |
| Religion |  |  |  |  |  |  |  |  |
| Christianity (Protestant) | 156(63.67) | 89(36.33) | Ref |  |  |  |  |  |
| Christianity (Catholic) | 87(67.44) | 42(32.56) | 0.84 | (0.53-1.32) | 0.468 |  |  |  |
| Muslim | 12(63.16) | 7(36.84) | 1.02 | (0.38-2.69) | 0.964 |  |  |  |
| None | 5(83.33) | 1(16.67) | 0.35 | (0.04-3.04) | 0.342 |  |  |  |
| Education level |  |  |  |  |  |  |  |  |
| None | 3(33.33) | 6(66.67) | Ref |  |  | Ref |  |  |
| Lower Primary | 12(92.31) | 1(7.69) | 0.04 | (0.003-0.49) | 0.012 | 0.06 | (0.004-0.92) | 0.044 |
| Upper Primary | 124(85.52) | 21(14.48) | 0.08 | (0.01-0.36) | 0.001 | 0.21 | (0.04-1.04) | 0.057 |
| Secondary | 106(53.27) | 93(46.73) | 0.43 | (0.10-1.80) | 0.253 | 0.89 | (0.19-4.14) | 0.884 |
| College/University | 15(45.45) | 18(54.55) | 0.6 | (0.12-2.81) | 0.517 | 0.76 | (0.14-4.06) | 0.757 |
| Marital Status |  |  |  |  |  |  |  |  |
| Single/living with parents/guardian | 222(71.84) | 87(28.16) | Ref |  |  | Ref |  |  |
| Married and living with partner | 16(64) | 9(36) | 1.43 | (0.61-3.36) | 0.407 | 1.32 | (0.43-4.00) | 0.615 |
| Single living alone | 19(32.76) | 39(67.24) | 5.23 | (2.86-9.56) | <0.0001 | 3.61 | (1.63-8.03) | 0.002 |
| Married not living with partner | 3(42.86) | 4(57.14) | 3.4 | (0.74-15.51) | 0.114 | 4.34 | (0.62-30.01) | 0.136 |
| Occupation |  |  |  |  |  |  |  |  |
| None | 49(56.32) | 38(43.68) | Ref |  |  | Ref |  |  |
| Student | 188(69.89) | 81(30.11) | 0.55 | (0.33-0.91) | 0.021 | 1.13 | (0.60-2.16) | 0.69 |
| Self employed | 17(48.57) | 18(51.43) | 1.36 | (0.62-2.99) | 0.438 | 0.74 | (0.29-1.87) | 0.532 |
| Employed | 6(75) | 2(25) | 0.42 | (0.08-2.25) | 0.317 | 0.17 | (0.02-1.07) | 0.061 |

### Summary of qualitative themes

Thematic analysis of FGDs and KIIs identified five cross-cutting themes. Limited knowledge and misuse of EC, sources of EC, perceptions of side effects, the influence of male partners, and unavailability of EC in public facilities, alongside a shared call for community-level education (Table 6)

**Table 6.** Summary of qualitative themes from FGDs and KIIs.

| Theme | FGDs (Adolescent Girls) | KIIs (Nurses) | KIIs (Chemists) |
| --- | --- | --- | --- |
| Limited Knowledge & misuse of ECs | Limited understanding of P2, with misconceptions about its use such as infertility and reproductive issues | Adolescents have inadequate knowledge on ECs; misuse it as a primary contraceptive method. | Adolescents misuse P2, using it frequently, instead of for emergencies. |
| Sources of Emergency Contraceptives | Mostly accessed through pharmacies or male partners, with public facilities rarely mentioned | Stock outs at public facilities, push clients to chemists for ECs. | High demand especially during holidays |
| Perceptions of Side Effects | Fears of infertility, cancer, and reproductive health problems | Myths around IUDs, leading to low uptake among adolescents. | Misconceptions persist; clients fear frequent use of P2 might cause harm. |
| Male Partner Influence | Male partners often provide P2, limiting girls' autonomy in contraceptive use. | - | Men occasionally buy P2 for their partners but are fewer than female clients. |
| Unavailability of ECs | - | Stock outs common in public facilities; clients referred to chemists. | Rapid stock turnover in chemists due to high demand, especially during school holidays |
| Community education needs | - | Need for community awareness campaigns on ECs use. | Community education on safe use needed. |

## DISCUSION

### Prevalence of emergency contraceptive use

The study finding showed that approximately one-third of the individuals surveyed (34.8%) admitted to using ECs. This percentage is slightly lower compared to the 43.5% reported among women of reproductive age in Siaya County during the 2022 Kenya Demographic Health Survey. The variance in numbers could be attributed to the fact the study specifically targeted populations, as our study focused on adolescents. The data from Kenya highlighted a concerning trend of a significant number of sexually active adolescents with low utilization rate of contraceptive services (Ohlendorf and Fehring 2007).

The study also revealed varying levels of EC uptake among adolescent girls, with use rates differing according to reproductive history. Adolescents with a history of pregnancy showed higher EC use rates than those without, though this difference was not statistically significant (p=0.083). Adolescents with children showed a significant association with higher EC uptake, especially those with one or two children (p=0.044). This finding could suggest a greater perceived need for EC use among girls with a history of childbirth, possibly to avoid additional pregnancies. Despite its common use, around 30%-50%, young people under 30 years have used ECs at a point in their lives. The use pattern is related to its availability and accessibility in combination with individual knowledge and awareness of ECs, with several barriers existing for women to utilize them effectively and consistently (Leon-Larios et al. 2022).

Findings from the KIIs, indicated a high demand for ECs among adolescents as reported by chemists. Sometimes it was used as a routine contraception rather than for emergencies. This indicates a lack of awareness of proper use. This suggests a need for better education on ECs and family planning methods. Emergency contraceptive use is not effective as a routine family planning method; it is recommended to use after unprotected sex to reduce chances of unwanted pregnancies. It is also effective when barrier methods such as slip-on methods fail or when intercourse was unpredictable (Kwame et al. 2022). However, although frequent use of emergency contraceptives is not recommended, repeated use does not pose significant health risks and should never be stated as a reason to deny women access (Mashoto et al. 2024).

### Access to and sources of emergency contraceptives

Most participants identified local chemists as their primary source of ECs, as few get from healthcare facilities. FGD participants emphasized chemists or male partners as primary sources, indicating potential barriers to accessing ECs from formal healthcare channels. KIIs with healthcare providers confirmed these findings, noting consistent stock outs of ECs in government health facilities, leading adolescents to rely on private pharmacies. However, study in Kenya indicated that emergency contraceptive pills are widely available in public clinics. They are part of post-rape care services. They are also commonly found in Kenya’s private sector, pharmacies, and clinics (Morgan et al. 2014).

This preference for pharmacies over healthcare facilities may be influenced by accessibility and perceived anonymity, as suggested by a key informant who noted low uptake of ECs even when they were in stock at facilities. Pharmtechs observed higher EC sales during school holidays, aligning with behavioral patterns of increased sexual activity during these periods. Many adolescent girls are at elevated risk of unintended pregnancy due to limited understanding of contraception, inadequate access to family planning services, or discomfort with using them. Furthermore, adolescents’ access to and awareness of regular contraception are low, and EC awareness is even lower, with very limited knowledge of the correct use of the method (Simmons et al. 2022). Addressing stock availability issues in government facilities could increase formal health facility utilization, promoting safer and more regulated access to ECs.

### Knowledge and perception of emergency contraceptives

Although most participants (95.2%) were aware of contraceptive methods, slightly over half were specifically familiar with ECs (commonly known as “P2”). Knowledge gaps remain, with 44.6% uncertain about correct timing and some mistakenly believing ECs prevent STIs. Misconceptions, such as frequent use causing infertility or cancer, may discourage effective use. Despite this, majority of users reported adhering to the recommended 72-hour window. According to Kenya Demographic Health Survey report of 2022, EC knowledge is at 33.4% among all women and 41.2% among unmarried sexually active women. This corresponds to a study conducted in Syria in 2023, which stated that: the knowledge of EC was low (36.1%) with the internet and social media being the primary sources of information (Alibrahim et al. 2024).

### Determinants of emergency contraceptive use

Unplanned sexual encounters, such as rape or condom failure, heavily drive EC use among adolescents. Qualitative data confirms that youth frequently visit public clinics following sexual violence, highlighting EC’s vital role in urgent situations.

To address this, healthcare facilities must integrate EC into broader sexual health services. Combining access with post-rape care, counseling, and emotional support creates a comprehensive continuum of care that protects vulnerable survivors.

The bivariate analysis revealed demographic factors heavily drive EC use among adolescents. Girls aged 15–17, those who are single, and those living alone show the highest likelihood of use. Conversely, older adolescents (18–19) and those with lower primary education are less likely to use EC.

In multivariate analysis, education level and marital status remained significant predictors of EC use, with adolescent girls who were single and living independently demonstrating higher odds of use (AOR: 3.61, p=0.002). This is confirmed by a study that linked higher education and having two or more children to high uptake of EC. High economic status has also been associated with contraceptive uptake (Atoyebi et al. 2025). The lower use rates among students may reflect a lack of information or a perception that ECs are unnecessary. By greater autonomy are more likely to make independent contraceptive decisions. Sexual health education, particularly for in-school adolescents, is critical to supporting safe and informed choices.

Reproductive history emerged as a key determinant, with adolescents having two children showing a significantly higher likelihood of EC use. Additionally, knowledge emerged as a substantial predictor. Adolescents aware of ECs or the specific “P2” pill were significantly more likely to use ECs. Conversely, those who held the misconception that ECs could be used effectively beyond 120 hours after intercourse were less likely to use them appropriately.

Several studies have reported limitations in the knowledge of respondents on the effective use of the emergency contraceptives. This is associated with the fact that the common source of information are friends where a times gives wrong information, misguides or having some misconceptions, thereby negatively affecting the choices to be made on which ECs to consider. Studies conducted in Kenya found that those who did not intend to use contraception most commonly cited fear of side effects and health concerns. However, it was also reported that even when the awareness is high, poor knowledge of contraceptive method and their side effects are associated with poor uptake. This is related to the myths and misconception that many women hold about potential side effects and negative outcomes (Ochako et al. 2015).

These myths are heard mostly from peers and partners who have a great influence on contraceptive demand and uptake (Tenaw, 2022). The overall knowledge of ECs has been reported to be high with low usage rate. This is justified by a study conducted by Mustapha which found the knowledge of EC to be at 90.6% while only 9.4% had good knowledge of ECs and its utilization (Hallidu and Sumaila 2022). It is crucial to stress the importance of timely use of ECs. It should be used within the 72-hour window and at the same time, prevent misuse.

This study revealed that adolescent EC use is influenced by reproductive history, sociodemographic factors, and knowledge gaps, emphasizing the need for targeted education, improved healthcare access, and addressing misconceptions about ECs to support informed, safe contraceptive use among adolescents. The WHO defines EC as a method of contraception that can prevent pregnancy after few days of intercourse. It is therefore intended for emergency use only following cases of unprotected sexual intercourse, failure or misuse of a contraceptive, coerced sex or rape. They are only effective in the first 120 hours following intercourse, before fertilization of the ovum (Demissie et al. 2020). The Family Health International (FHI) report of November 2021 stated that ECs are more effective when started soon after unprotected intercourse. The report further stated that the progestin-only pills prevent about 95% of pregnancies if started within 24 hours of unprotected intercourse, 85% for initiation in the 25^th^ to 48^th^ hours, and 58% when started in 49^th^ to 72^nd^ hours. EC is recommended within five days after sexual intercourse though it is more effective when used as soon as possible when it can prevent an unwanted pregnancy but cannot cause an abortion or damage a developing embryo.

The EC is a valuable option that should not be relied upon as a primary method of contraception and does not provide protection against sexually transmitted diseases. (Jiménez-Iglesias et al. 2018). When used after unprotected sexual intercourse, emergency contraceptive pills can help reduce the risk of pregnancy. These pills are intended for occasional use and are not as effective as other hormonal or long-acting contraceptive methods, so they are not recommended for regular use. There is no consensus on the frequency of EC pill use, but the World Health Organization suggest a limit of 1-4 times per month based on a study of efficacy and side effects of repeat use(Facultad de Medicina, Universidad Cooperativa de Colombia. Villavicencio (Colombia) et al. 2022). It is important to remember that is just one EC tool in a comprehensive approach to sexual health and family planning

### Strengths and limitations

This study utilized a convergent mixed-methods design to combine quantitative data with qualitative insights on adolescent emergency contraception (EC) use. While triangulation helped mitigate potential recall and social-desirability biases inherent in self-reported data, the cross-sectional approach prevents causal inference. Additionally, because the findings focus exclusively on a single rural ward, caution is required when generalizing the results to diverse urban or peri-urban populations.

## CONCLUSION

EC use among adolescents aged 15–19 years in West Asembo Ward was 34.8% despite high awareness. Uptake was associated with age, marital status, independent living, and parity. Stock-outs and privacy concerns in public facilities encouraged adolescents to obtain EC from private pharmacies. The County Ministry of Health should strengthen adolescent-friendly services, ensure consistent EC availability, and provide confidential, non-judgmental care. Targeted school and community education should address misconceptions and promote appropriate EC use, particularly among young mothers and adolescents with limited education.

## DECLARATIONS

### Availability of data and materials

The datasets generated and/or analysed during the current study are available from the corresponding author on reasonable request.

### Authors’ contributions

GOA conceived and designed the study, led data collection, analysed the data, and drafted the manuscript. DO, SO and IO supervised the study design and analysis and critically revised the manuscript for important intellectual content. All authors read and approved the final manuscript.

## Data Availability

All data produced in this work are contained in the manuscript

https://www.george.com

## Acknowledgements

The authors thank the adolescent girls, healthcare providers, and chemist attendants of West Asembo Ward who participated in this study, the community health promoters who supported household enumeration and mobilization, and the field assistants who supported data collection.

